# The polymorphism L412F in *TLR3* inhibits autophagy and is a marker of severe COVID-19 in males

**DOI:** 10.1101/2021.03.23.21254158

**Authors:** Susanna Croci, Mary Anna Venneri, Stefania Mantovani, Chiara Fallerini, Elisa Benetti, Nicola Picchiotti, Federica Campolo, Francesco Imperatore, Maria Palmieri, Sergio Daga, Chiara Gabbi, Francesca Montagnani, Giada Beligni, Ticiana D.J. Farias, Miriam Lucia Carriero, Laura Di Sarno, Diana Alaverdian, Sigrid Aslaksen, Maria Vittoria Cubellis, Ottavia Spiga, Margherita Baldassarri, Francesca Fava, Paul J. Norman, Elisa Frullanti, Andrea M. Isidori, Antonio Amoroso, Francesca Mari, Simone Furini, Mario U Mondelli, GEN-COVID multicenter study, Mario Chiariello, Alessandra Renieri, Ilaria Meloni

**Author notes:** **Corresponding author:** Professor Alessandra Renieri, Medical Genetics Unit, University of Siena, Policlinico Le Scotte, Viale Bracci, 2, 53100 Siena, Italy, E.mail, Dr. Mario Chiariello, Core Research Laboratory (CRL), Institute for the Study, Prevention and Oncology Network (ISPRO), Via Fiorentina 1, 53100 Siena, E.mail.

## Abstract

The polymorphism L412F in *TLR3* has been associated with several infectious diseases. However, the mechanism underlying this association is still unexplored. Here, we show that the L412F polymorphism in *TLR3* is a marker of severity in COVID-19. This association increases in the sub-cohort of males. Impaired autophagy and reduced TNFα production was demonstrated in HEK293 cells transfected with TLR3-L412F plasmid and stimulated with specific agonist poly(I:C). A statistically significant reduced survival at 28 days was shown in L412F COVID-19 patients treated with the autophagy-inhibitor hydroxychloroquine (P=0.038). An increased frequency of autoimmune disorders as co-morbidity was found in L412F COVID-19 males with specific class II HLA haplotypes prone to autoantigen presentation. Our analyses indicate that L412F polymorphism makes males at risk of severe COVID-19 and provides a rationale for reinterpreting clinical trials considering autophagy pathways.

## INTRODUCTION

In December 2019, a new virus was isolated in Wuhan, China, which was called Severe Acute Respiratory Syndrome Coronavirus 2 (SARS-CoV-2). SARS-CoV-2 is an enveloped positive-sense RNA virus that caused a new pandemic which WHO named COVID-19 (CoronaVirus Disease-2019).

To date, many characteristics of SARS-CoV-2 are still unclear and, although its ability to be transmitted from one person to another has been ascertained, uncertainties remain about the exact modes of transmission and pathogenicity. In addition, a high variability of symptoms in infected patients and between different populations has been reported; one of the possible explanations of such variability is the genetic background of the host that may affect immune responses to the virus. Among host genetic factors that might impact on symptoms severity there are genes involved in virus entry and mediators of innate immunity [1,2]. Toll-Like Receptors (TLRs) are a class of proteins that play a key role in host innate immunity, causing the production of pro-inflammatory cytokines (TNF-α, IL-1, and IL-6) and type I and II Interferons (IFN), that are responsible for innate antiviral responses. Among TLR genes, TLR3 is an interferon-inducing dsRNA sensor, whose activation is involved in protection against different RNA viruses [2-4]. Upon viral infection, TLR3 signalling leads to the activation of two factors, NF-kB and Interferon (IFN)-Regulatory Factor 3 (IRF3), that play an essential role in the immune response. This results in the production of various cytokines, including tumor necrosis factor-α (TNF-α), activating immune responses. However, increased inflammatory responses can make the patient more susceptible to pneumonia and autoimmune diseases. Accordingly, a protective effect against fatal pneumonia has been reported in the absence of TLR3 [5-7]. Among TLR3 variants, the functional L412F polymorphism (rs3775291; c.1234C>T) is known to decrease TLR3 expression on the cell surface [8]. This polymorphism also leads to poor recognition of SARS-CoV-2 dsRNA compared to its wild type counterpart [9] and has been recently associated with SARS-CoV-2 susceptibility and mortality [10].

There is evidence that TLR3, as other TLRs, acts through autophagy in determining susceptibility to infections [11]. The autophagic pathway is essential during infection and for molecular processes as cell maintenance and homeostasis [11-14]. Indeed, autophagy is one of the major cell defence mechanisms against pathogens [15]. A role for autophagy is reported in different studies on other Coronaviruses such as the mouse hepatitis virus (MHV) and the transmissible gastroenteritis virus (TGEV) [16,17]. A role in SARS-CoV-2 infection has also been described [18-20]. In particular, SARS-CoV-2 can inhibit autophagy resulting in accumulation of autophagosomes and inhibition of viral clearance that, together with immune dysfunction and the activation of numerous inflammatory cytokines, leads to a more severe form of COVID-19 [21-23].

To shed light on the mechanisms underlying the diverse susceptibility to COVID-19, we performed a nested-control study within our GEN-COVID cohort, confirming the role of L412F polymorphism in *TLR3* gene in susceptibility to SARS-CoV-2 and further defining the potential mechanisms by which this effect is exerted.

## RESULTS AND DISCUSSION

Comparing the extreme phenotypes of SARS-CoV-2 infection, severe COVID-19 patients (cases) versus SARS-CoV-2 PCR-positive oligo-asymptomatic subjects (controls), and using LASSO Logistic regression on common bi-allelic polymorphisms from whole-exome sequencing, we identified the L412F polymorphism (rs3775291; c.1234C>T) in *TLR3* as a severity marker (**Fig. 1A**). The grid search curve of the cross-validation score (**Fig.1B**) shows a maximum of the regularization parameter in 10. With this calibration setting, the 10-fold cross-validation provides good performances in terms of accuracy (73%), precision (74%), sensitivity (73%), and specificity (73%) as shown in **Fig.1C**. The confusion matrix is reported in **Fig.1D**, whereas the Receiver Operating Characteristic (ROC) curve (**Fig.1E**) provides an Area Under the Curve (AUC) score of 80%.

**Fig.1.**
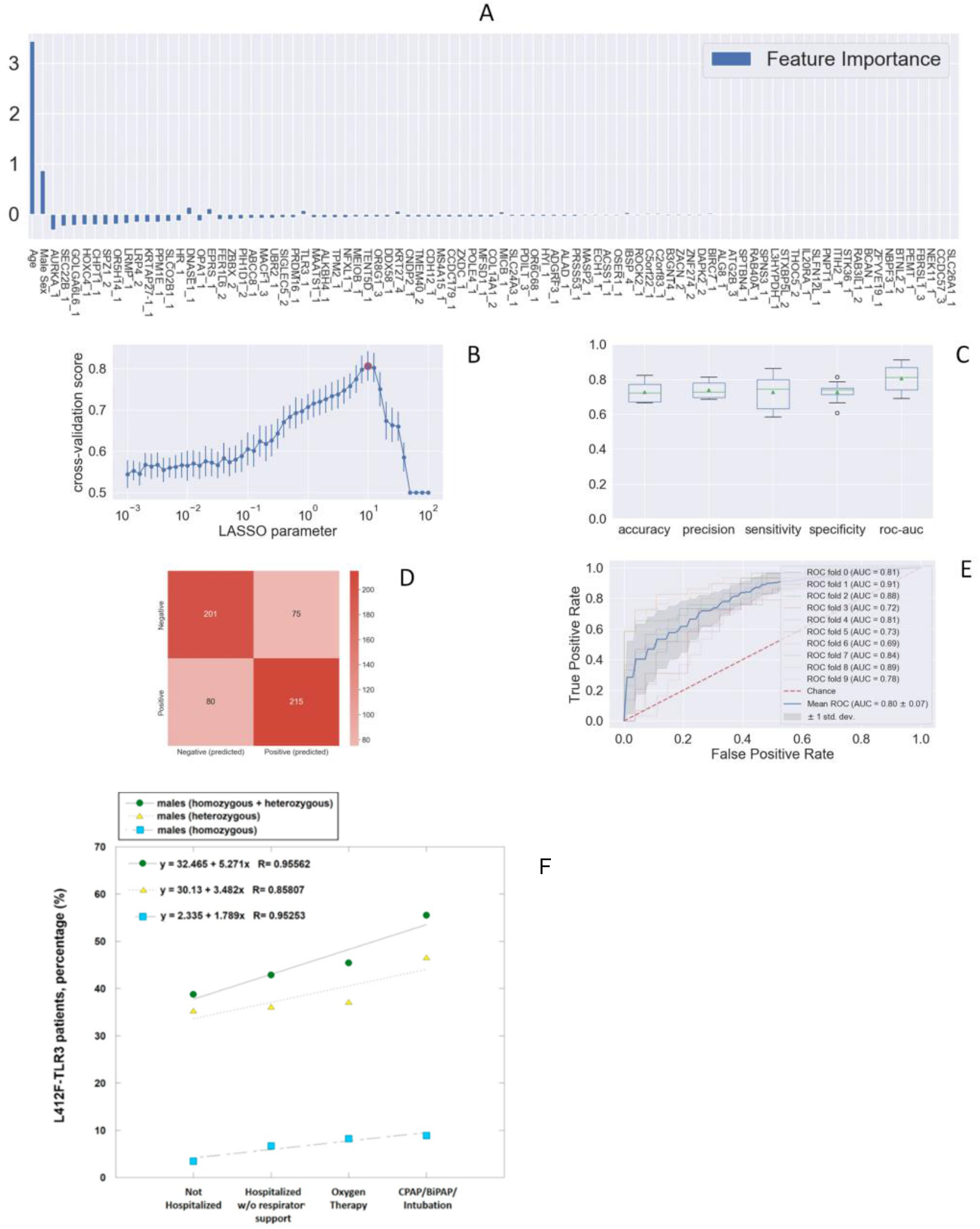
The histogram of the LASSO logistic regression weights represents the importance of each feature for the classification task, **Panel A**. The positive weights reflect a susceptible behaviour of the features to the target COVID-19 disease, whereas the negative weights a protective action. **Panel B**: Cross-validation ROC-AUC score for the grid of LASSO regularization parameters; the error bar is given by the standard deviation of the score within the 10 folds; the optimal regularization parameter is chosen by selecting the one with highest cross-validation score (red point). **Panel C**: Boxplot of accuracy, precision, sensitivity, specificity, and ROC-AUC score for the 10-fold of the cross-validation. The box extends from the Q1 to Q3 quartile, with a line at the median (Q2) and a triangle for the average. **Panel D**: Confusion matrix for the aggregation of the logistic regression predictions in the 10 folds of the cross-validation. **Panel E**: ROC curve for the 10 folds of the cross-validation. **Panel F**. Distribution of carriers of the polymorphism L412F in homozygous or heterozygous states stratified by clinical category.

The L412F polymorphism has an overall allele frequency of about 20%, ranging from 30% in European to 0.88% in African (mainly sub Saharan) populations [8]. It is intriguing that a COVID-19-free population such as sub Saharan has a very low frequency (0.88%) of this polymorphism and that Asian (26.97%) and European (30.01%) have a much higher frequency. The variant protein with phenylalanine is under-represented on the cell surface, it is not efficiently secreted into the culture medium when expressed as the soluble ectodomain, and it has reduced capability to activate the expression of TLR3-dependent reporter constructs [8]. In order to confirm the role of the polymorphism, we compared individuals showing severe COVID-19 (cases) and those with no sign of the disease (controls). We subdivided patients in two categories, those having the polymorphism in heterozygous or homozygous state and those homozygous for the wild type allele. We found that the prevalence of L412F polymorphism is significantly higher in cases compared to controls (p-value 2.8×10^−2^) (**Table 1a**). The global allele frequency of L412F in our cohort (cases and controls) is 29.38%, like to the allele frequency of 29.79% reported in the European (non-Finnish) population in the gnomAD database (https://gnomad.broadinstitute.org/). The identified frequencies were in Hardy-Weinberg equilibrium.

**Table 1a.**
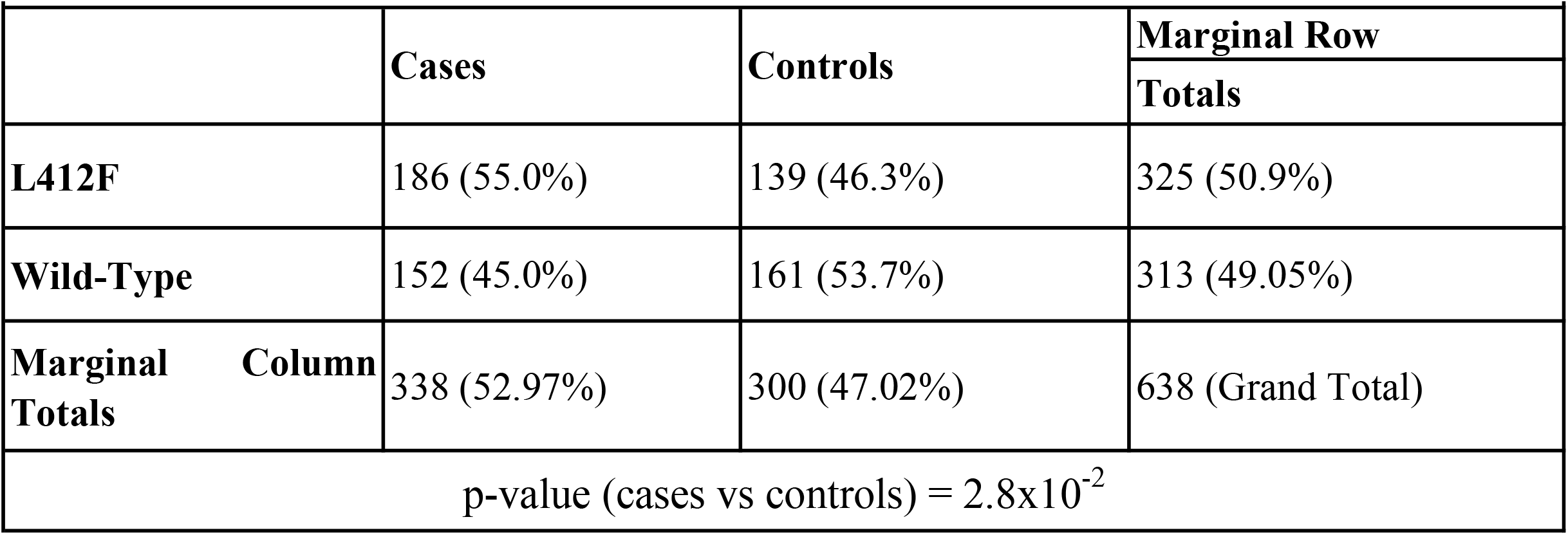
L412F and COVID-19 outcome (both sexes)

**Table 1b.**
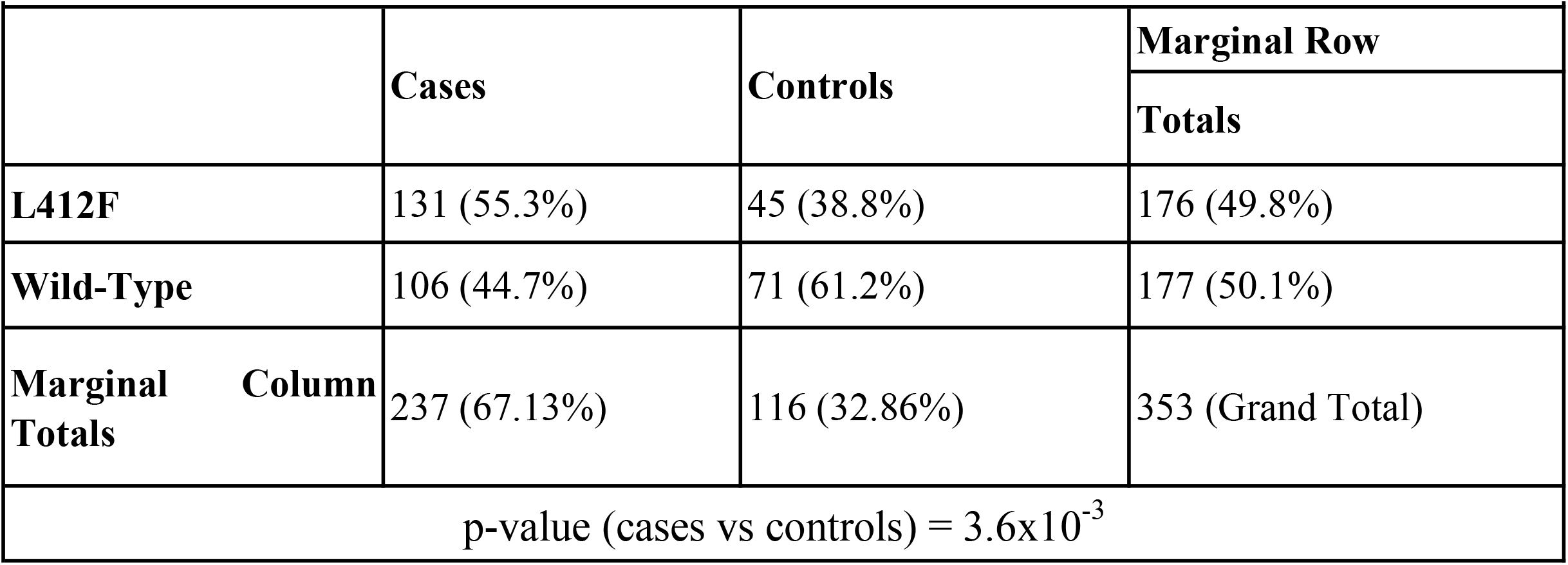
L412F and COVID-19 outcome (males only)

**Table 1c.**
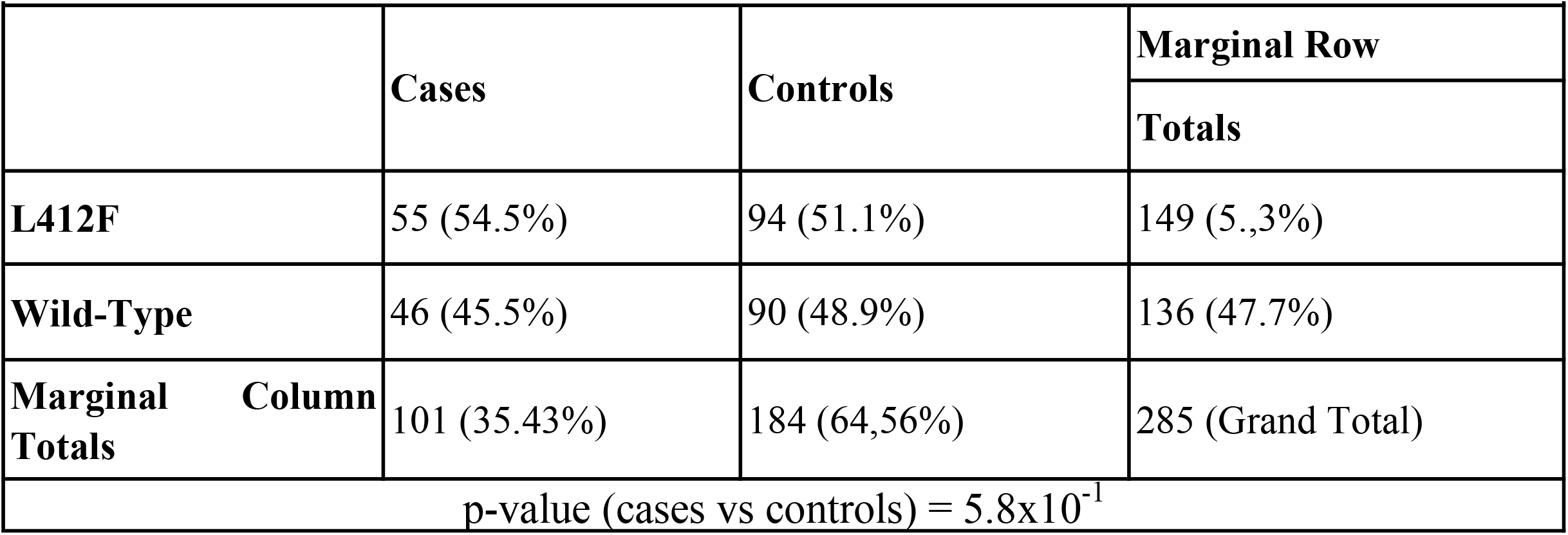
L412F and COVID-19 outcome (females only)

Sex-related differences of TLRs activation following stimulation by viral nucleic acid may be involved in the sex-related variability in response to viral infections [24]. Several rare TLR3 loss of function mutations are known linked both to influenza and SARS-CoV-2 virus as well as hyperfunctioning mutations [25, 26]. In agreement with these data, when we stratified by gender, the statistically significant difference increased in the sub-cohort of males giving an OR of 1.94 (95% CI, 1.23 to 3.06; p=3.8×10^−3^), while it was lost in the sub-cohort of females (p-value 5.8×10^−1^), (**Table 1**).

We then investigated the prevalence of patients carrying L412F in heterozygous or homozygous states in all 4 categories of COVID-19 clinical severity, considering only male subjects regardless of age (n=665). We found that the prevalence of carriers directly increased with the severity of COVID-19, from a clinical condition not-requiring hospitalization to intratracheal intubation (**Fig. 1F**)

The L412F substitution in TLR3 falls in the ectodomain, in the 14 Leucine-Rich Repeats (LRR) domain, a motif of 22 amino acids in length that folds into a horseshoe shape [27]. Proteins containing LRRs are involved in a variety of biological processes, including signal transduction, cell adhesion, DNA repair, recombination, transcription, RNA processing, disease resistance, apoptosis, and the immune response [slit: an extracellular protein necessary for development of midline glia and commissural axon pathways contains both EGF and LRR domains [28]. The L412F substitution is expected to have a limited structural impact with minimal rearrangement of near hydrophobic amino acids like tryptophan 386 (**Fig. 2**). However, the absence of one of the leucines probably determines a different rearrangement of the motif and consequently of the near glycosylation site ASN414, having an impact on protein-protein interaction and in signal transduction process [29].

**Figure 2.**
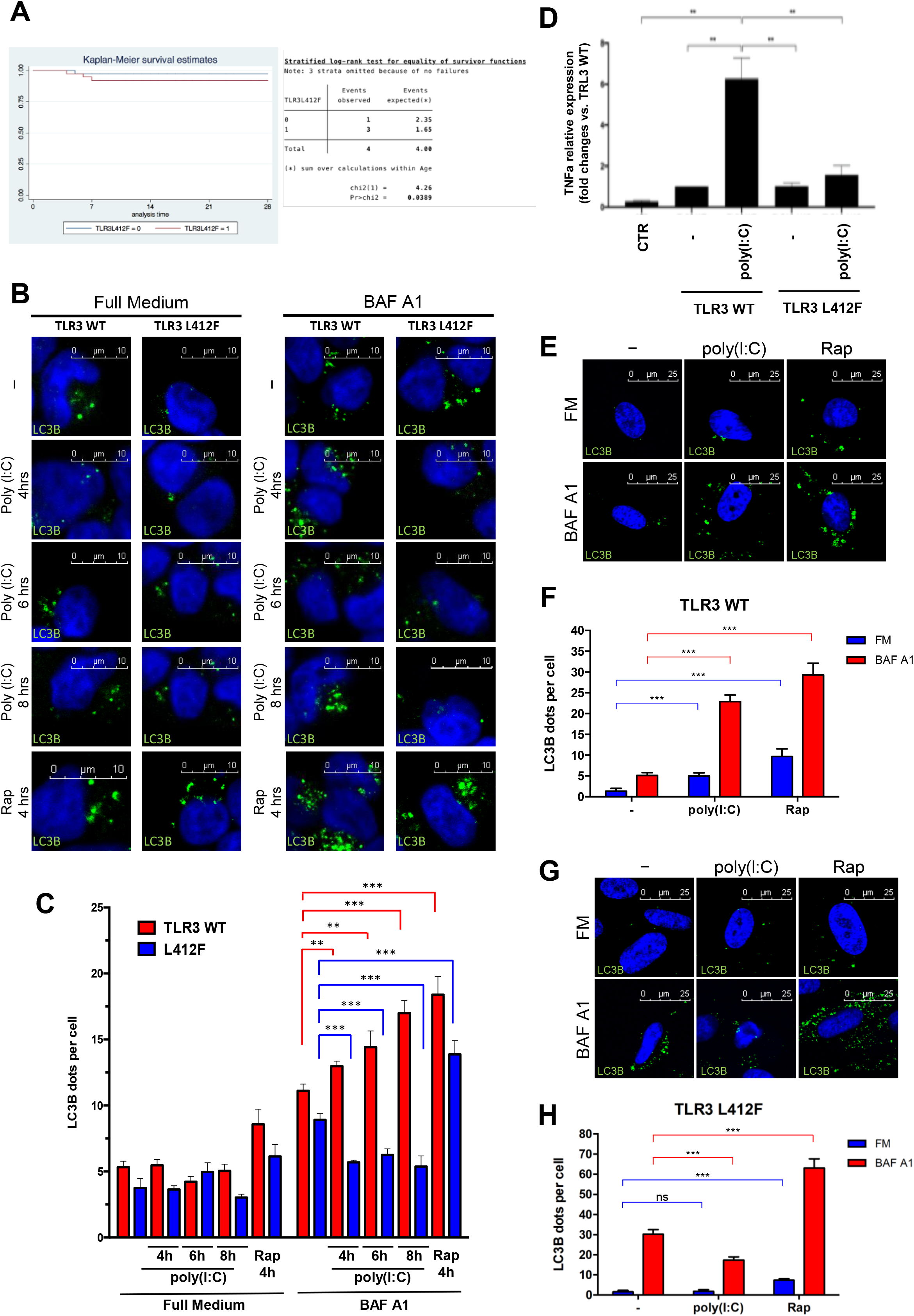
Superposition of wild type and mutated TLR3 protein. **Panel A**: TRL3 human protein tridimensional structure of 2Z7X crystal structure. In green cartoon representation of TLR3 protein. In **Panel B** and **Panel C** Zoom of mutated region with LEU412 in red sticks and PHE412 in magenta. The hydrophobic core of Leu377, Leu389, and Trp386 in blue sticks.

Germline knockout of *TLR3* inhibits autophagy and upregulation of TLR3 promotes damage after myocardial infarction mainly because of autophagy rather than inflammatory activation [30]. Interestingly, we could notice a statistically significant (p=3.8×10^−2^) reduced survival at 28 days in TLR3_L412F COVID-19 patients treated with hydroxychloroquine (HCQ) (**Fig.3A)**. As this drug is a well-established inhibitor of autophagy, we reasoned that alterations of this important biological process might have a role in the increased severity of the clinical phenotypes of SARS-CoV-2 infection in patients with the TLR3_L412F polymorphism. Notably, beside being entirely ineffective at changing the clinical evolution of COVID-19, which led to retraction of the paper reporting the clinical trial [31], HCQ may have been responsible for reportedly increased fatality rates among patients treated with this drug [32,33]. Poly(I:C) stimulation of the TLR3 receptor has already been shown to stimulate autophagy [30]. Therefore, we decided to compare the efficacy of transfected WT and L412F-mutated receptors in inducing autophagy upon poly(I:C) treatment. To monitor autophagy, we used indirect immunofluorescence microscopy to score the formation of punctate intracellular vacuoles stained for LC3B. Moreover, to better appreciate autophagosomal formation, we also used bafilomycin A1 (Baf A1), an inhibitor of lysosomal acidification, to prevent lysosomal degradation of autophagosome-associated LC3B [34]. Baf A1 allows, in fact, the detection of each autophagosome formed in the time-lapse between addition of the drug to cells and harvesting [34]. Importantly, to avoid potentially confounding effects of the endogenous TLR3 receptor in transfected cells, we used TLR3 knock-out HEK cells. In these cells, when transfected with TLR3_WT, we observed a progressively increasing number of autophagosomes (AP) when stimulating with poly(I:C) for different time points in the presence of BAF A1 (compared to BAF A1 alone) (**Fig. 3B-C**), indicating a stimulation of the synthesis of these vesicles and a positive autophagic flux. Conversely, in HEK cells transfected with TLR3_L412F, the number of AP was reduced by poly(I:C) stimulation in the presence of BAF A1 (**Fig. 3B-C**), demonstrating a block in AP synthesis and a reduced autophagic flux. Interestingly, in the absence of BAF A1, AP numbers did not increase upon poly(I:C) stimulation, suggesting that, in HEK cells, fast degradation of AP may compensate for a small increase in the synthesis. Indeed, also rapamycin (RAP), a strong stimulus for autophagy [35], induced only a small increase of AP in the absence of BAF A1 in these cells upon transfection of both TLR3_WT and TLR3_L412F, supporting slow rates of AP synthesis in these cells upon stimulation with different stimuli.

**Fig. 3.**
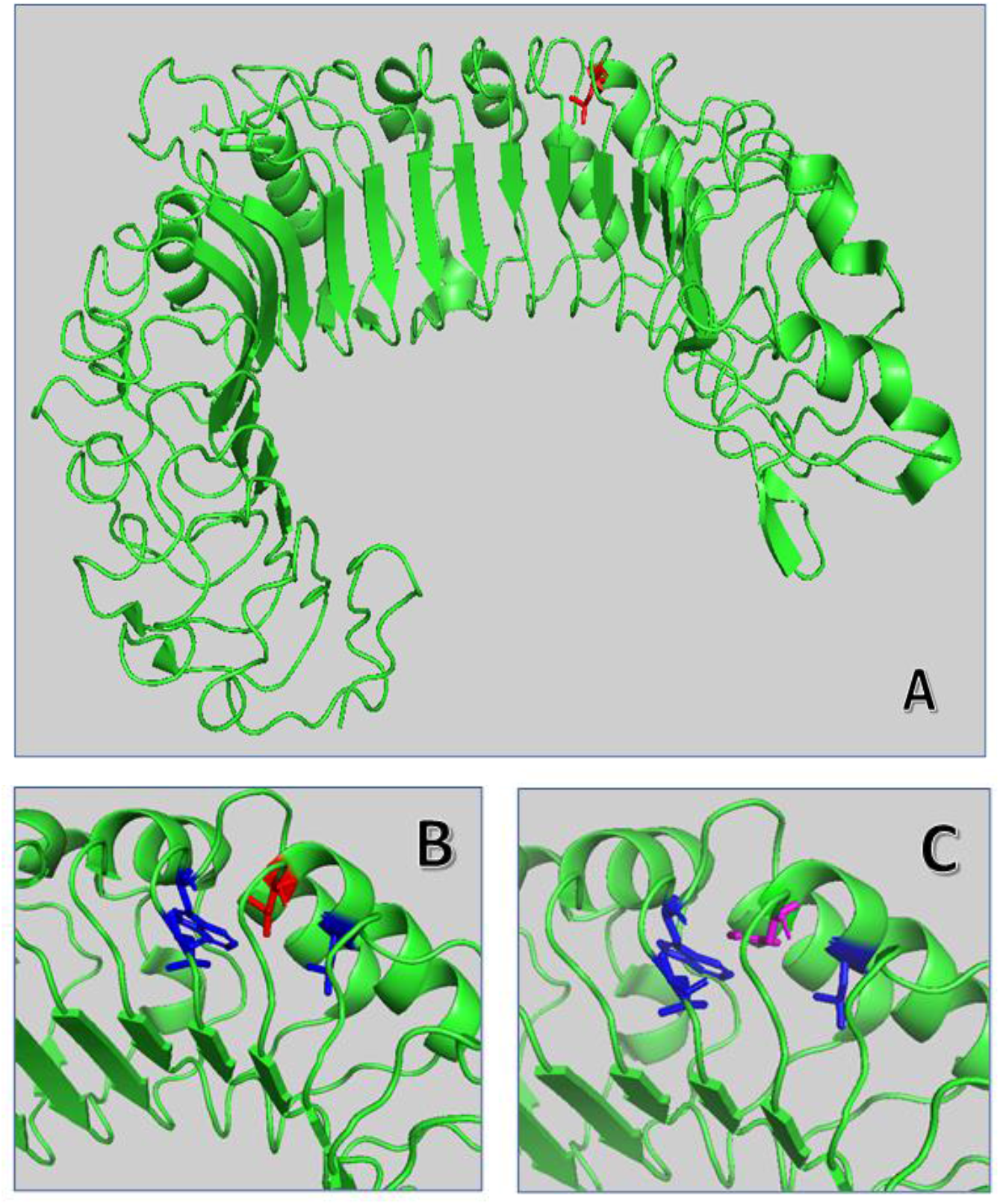
Analysis of autophagy in TLR3_L412F expressing cells. **Panel A**: 28-day survival study of TLR3-L412F carriers vs not-carriers in the group treated with hydroxychloroquine. N=156, with 73 carriers of TLR3-L412F. 3 carriers and 1 not-carrier died in the first 28 days of treatment. **Panel B:** Analysis of autophagy in HEK-KO cells expressing wild type or L412F mutant proteins. HEK-KO cells were transfected for 24 hrs with plasmids encoding TLR3_WT and TLR3_L412F. Cells were next incubated in full medium (FM) or FM + 400 nM Bafalomycin A1 (BAF A1) for 3 hrs and stimulated for increasing times with 50 µg/ml Poly(I:C), as indicated. Cells were next fixed, permeabilized with 100 µg/ml digitonin and stained with anti-LC3B antibodies and revealed with AlexaFluor488-conjugated secondary antibodies. Nuclei were stained with DAPI. Where indicated, rapamycin (RAP, 500 nM, for 2 hrs) was used as positive control for induction of autophagy. **Panel C**: Same as in B, but the amount of autophagosomes (scored as LC3B-positive dots) per cell was quantified by Volocity software. Measures were obtained by analyzing at least 400 cells/sample from 3 different experiments (n = 3). **Panel D:** Analysis of TNF-α mRNA expression in HEK-KO cells expressing wild type or L412F mutant proteins. HEK-KO cells were transfected for 24 hrs with plasmids encoding empty vector (CTR), TLR3_WT and TLR3_L412F and next stimulated for increasing times with 50 µg/ml Poly(I:C), where indicated. TNF-α levels were evaluated by Real Time PCR. The gene expression levels were evaluated by the fold change versus TLR2 WT sample using the equation 2^-DDCt^. Data are presented as the mean ± SEM. Data significance was analyzed using One-way ANOVA test with Holm-Sidak’s correction. Asterisks were attributed for the following significance values: P > 0.05 (ns), P < 0.05 (*) and P < 0.01 (**). **Panel E:** Normal human fibroblasts (NDHF) from subjects expressing the TLR3_WT receptor were stimulated with 50 µg/ml Poly(I:C) or RAP (1 µ M) for 4 hrs, in full medium alone or containing 400 nM bafilomycin A1 for 3 h. Cells were next fixed, permeabilized with 100 µg/ml digitonin and stained with anti-LC3B antibodies and revealed with AlexaFluor488-conjugated secondary antibodies. Nuclei were stained with DAPI. **Panel F:** Same as in E, but the number of autophagosomes (scored as LC3B-positive dots) per cell was evaluated for each sample by Volocity software. G and H) same as in E-F, but fibroblasts are homozygous for the TLR3_L412F receptor. Statistical analysis was performed using Student’s t test. Means § SEM for each value are shown in the graphs. ns=not significant; ** = p < 0.01; *** = p < 0.001.

During SARS-CoV-2 infection, adipocytes produce pro-inflammatory cytokines like TNF-α, IL-6 and IL-1 which recruit immune cells to the site of infection [36]. Autophagy is stimulated and regulated by these pro-inflammatory cytokines [36]. TNF-α is a potent immunomodulator and proinflammatory cytokine that has been implicated in the pathogenesis of autoimmune and infectious diseases. It is produced by activated monocytes and macrophages, as well as by many other cell types, including lymphocytes, as a transmembrane protein. Through cell modulation, TNF-α can activate both cell death and survival mechanisms. TNF-α induces autophagy through a feedback mechanism, causing further recruitment and activation of lymphocytes and contributing to the excess inflammation typical of SARS-CoV-2 infection [37]. As chloroquine, a powerful inhibitor of autophagy, inhibits production of different cytokines, among which TNF-α [38], we next decided to test if the inhibitory effect of the L412F mutation on autophagy was also able to mimic the effect of the pharmacological inhibitor of this process in HEK cells. Indeed, while poly(I:C) readily stimulated TNFα expression in HEK cells transfected with the TLR3_WT receptor, this effect was completely abolished in TLR3_L412F transfected cells (**Fig. 3D**).

In order to validate data obtained on transfected HEK cells, we next isolated and cultured skin fibroblasts from healthy donors with different genotypes relative to the TLR3 locus: wild-type (WT/WT) and L412F (L412F/L412F) homozygous and WT/L412F heterozygous. In these primary fibroblasts, immunofluorescence analysis revealed that the number of LC3B-positive vesicles increased upon poly(I:C) stimulation both in the absence and in the presence of BAF A1 (**Fig. 3E-F**), showing an overall positive autophagic flux in these cells, while resulting significantly reduced in L412F/L412F fibroblasts (**Fig. 3G-H**). As a control, rapamycin stimulation of both WT/WT and L412F/L412F resulted in a positive autophagic flux (**Fig. 3E-F and G-H**) confirming that the mutation specifically affected TLR3-dependent autophagy and not the general autophagic process.

Overall, our results therefore suggest that the outcome of clinical trials with HCQ should be reinterpreted in the light of TLR-L412F polymorphism status. Negative effects of the drug in L412F bearing subjects, may have masked a possible positive outcome in L412F-free subjects. *TLR3* variant L412F has been associated with a wide range of autoimmune diseases including Addison’s disease and hypothyroidism [39].

*TLR3* rare variants resulting in partial loss of function and occurring together with the common variant L412F, or with another rare variant, have been identified in Addison’s disease [40]. Persistent viral infections in a background of defective innate immunity lead to overexpression of HLA haplotypes prone to present autoantigen. Defects of autophagy have been observed in many infectious and autoimmune diseases. Alteration of autophagic processes causes the onset of autoimmunity due to increased survival and reduced apoptosis of self-reactive lymphocytes [41-43]. HLA has been shown to be implicated in disease severity and clinical outcome of patients with COVID-19 [44]. Accordingly, an increased frequency of autoimmune disorders as co-morbidity was found in our cohort in L412F COVID-19 patients with specific HLA class II haplotypes prone to autoantigen presentation. In particular, we analysed a DR3-DQ2 haplotype which predisposes to different types of autoimmune diseases [45,46]. The frequency of autoimmune disorders is indeed significantly increased in male patients with HLA DR3/DQ2 haplotype and L412F, especially diabetes (25%) (**Table 2a and b**). These results suggest that the combination of L412F in *TLR3* and a specific class II HLA haplotype puts male patients at risk of post-COVID autoimmune exacerbation emphasizing the need for appropriate follow-up.

**Table 2a.**
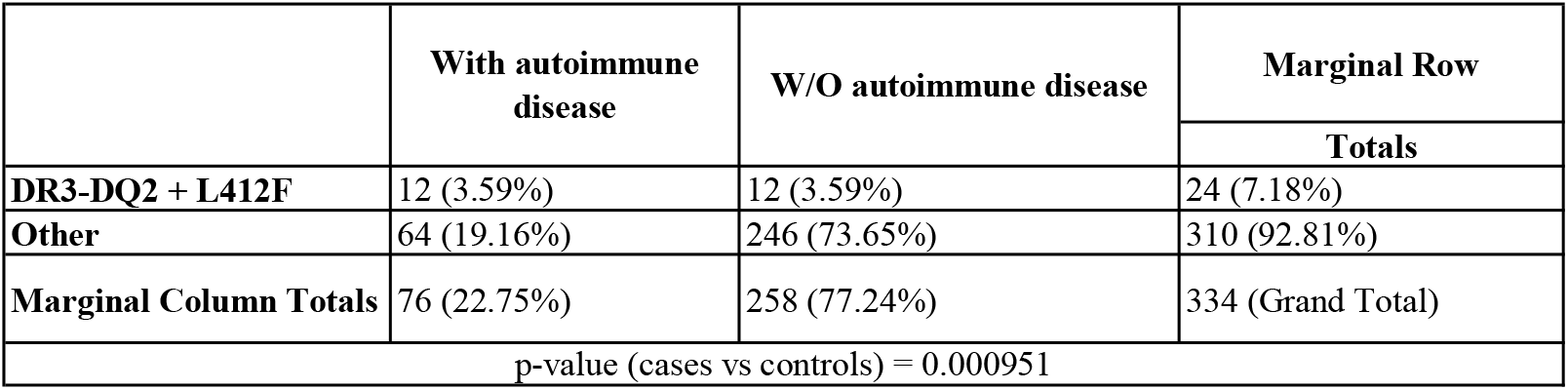
Association between DR3-DQ2 + L412F haplotype and autoimmune disorders in male patients

|  | With autoimmune disease | W/O autoimmune disease | Marginal Row |
| --- | --- | --- | --- |
|  |  |  | Totals |
| **DR3-DQ2 + L412F** | 12 (3.59%) | 12 (3.59%) | 24 (7.18%) |
| **Other** | 64 (19.16%) | 246 (73.65%) | 310 (92.81%) |
| **Marginal Column Totals** | 76 (22.75%) | 258 (77.24%) | 334 (Grand Total) |
| p-value (cases vs controls) = 0.000951 |  |  |  |

**Table 2b.** Male patients with L412F and HLA DR3/DQ2 haplotype

| PatientID | Age | Ethnicity (white=1, hispanic=4) | Clinical Category | GT TLR3 (L412F) | Clinical known comorbidities |
| --- | --- | --- | --- | --- | --- |
| AR-COV-25 | 69 | 1 | 4 | 0/1 | Hypertension, Kidney failure, COLD |
| BS-COV-102 | 59 | 1 | 3 | 0/1 | Diabetes Mellitus, diabetic neuropathy with transmetatarsal amputation, bilateral fasciectomy, left hernioplasty, diabetic retinopathy |
| BS-COV-58 | 47 | 1 | 3 | 0/1 | Hypertension |
| BS-COV-65 | 72 | 1 | 2 | 0/1 | Rheumatoid Arthritis |
| BS-COV-70 | 65 | 1 | 3 | 0/1 | Autoimmune hepatitis, autoimmune polyglandol syndrome, previous autoimmune thyroid |
| CR-COV-17 | 61 | 1 | 2 | 0/1 | Inflammatory disease, Hypertension |
| CR-COV-4 | 66 | 1 | 3 | 0/1 | None |
| LS-COV-10 | 37 | 1 | 1 | 0/1 | DLB-CL EBV+, Immunodeficiency, Bipolar Disorder |
| LS-COV-19 | 58 | 4 | 2 | 0/1 | Diabetes Mellitus |
| LS-COV-8 | 48 | 1 | 4 | 0/1 | Hypertension |
| MORE-COV-16 | 65 | 1 | 3 | 0/1 | Hypertension, Asthma, Dyslipidemia |
| MORE-COV-7 | 72 | 1 | 4 | 0/1 | Hypertension |
| PG-COV-15 | 87 | 1 | 2 | 0/1 | Congestive heart failure, Dyslipidemia, |
| PG-COV-23 | 68 | 1 | 1 | 0/1 | N/A |
| PV-COV-02 | 65 | 1 | 2 | 0/1 | Rheumatoid Arthritis |
| PV-COV-23 | 72 | 1 | 3 | 0/1 | Hypertension, Diabetes Mellitus |
| PV-COV-24 | 79 | 1 | 2 | 0/1 | Atrial Fibrillation, Diabetes Mellitus, Kidney failure |
| PV-COV-39 | 33 | 4 | 4 | 0/1 | Diabetes Mellitus, Obesity |
| PV-COV-73 | 54 | 1 | 3 | 0/1 | None |
| PV-COV-97 | 66 | 1 | 4 | 0/1 | Congestive Heart Failure, Hypertension, Diabetes Mellitus |
| RUF-COV-1 | 52 | 1 | 0 | 0/1 | None |
| SPC-COV-14 | 56 | 1 | 3 | 0/1 | Asthma, HCV |
| TV-COV-81 | 44 | 1 | 1 | 0/1 | None |
| TV-COV-96 | 51 | 1 | 2 | 0/1 | Hypertension |

No association was found between AIRE loss of function variants and COVID-19 outcome, as outlined by the absence of the gene in Figure 1.

In conclusion, we have identified the second protein-encoding polymorphism that modulates COVID-19 outcome. These results indicate that L412F polymorphism in the *TLR3* gene makes males, in whom after puberty testosterone lowers TLR3 expression, at risk of severe COVID-19 in a context of a polygenic model. Moreover, based on impairment of autophagy, these data provide a rationale for reinterpreting clinical trials with HCQ stratifying patients by L412F. Finally, the combination of L412F in *TLR3* and specific HLA class II haplotypes may put male patients at risk of post-acute sequelae of SARS-CoOV-2 infection (PASC) pointing to the need for an appropriate follow-up. Our experiments suggest an important role of autophagy downstream the TLR3 receptor, possibly affecting TNFa production and susceptibility to infections, including SARS-CoV-2.

## MATERIAL AND METHODS

### Patients

We performed a nested case-control study (NCC). We used a cohort of 1319 subjects (cases and controls) from the Italian GEN-COVID Multicenter study, infected with SARS-CoV-2 diagnosed by RT-PCR on nasopharyngeal swab [47]. Cases were defined as patients needing endotracheal intubation or CPAP/biPAP ventilation. Controls were oligo-asymptomatic subjects not requiring hospitalization.

### LASSO logistic regression

We adopted 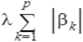, the LASSO logistic regression model for the classification of severe COVID-19 patients (cases) versus SARS-CoV-2 PCR-positive oligo-asymptomatic subjects (controls), able to enforce both the sparsity and the interpretability of the results. By denoting with *β*_□_ the coefficients of the logistic regression and by lambda (*λ*) the strength of the regularization, the LASSO (Least Absolute Shrinkage and Selection Operator) regularization [48] term of the loss, has the effect of shrinking the estimated coefficients to 0. In this way, the weights of the logistic regression algorithm can be interpreted as the feature importances of the subset of the most relevant features for the task [49]. The input features are the common bi-allelic polymorphisms from whole-exome sequencing as well as gender, and the age, the latter as a continuous variable normalized between 0 and 1. Common bi-allelic polymorphisms are defined as combinations of two polymorphisms, each with MAF above 1%, with frequency above 5% in the cohort.

The fundamental hyper-parameter of the logistic regression algorithm is the strength of the LASSO term, which is tuned with a grid search method on the average area under the Receiver Operating Characteristic (ROC) curve for the 10-fold cross-validation. The regularization hyperparameter varies in the range [10^−3^, 10^2^] with 50 equally spaced values in the logarithmic scale. The optimal regularization parameter is chosen by selecting the parameter with the highest cross-validation score. During the fitting procedure, the class slight unbalancing is tackled by penalizing the misclassification of the minority class with a multiplicative factor inversely proportional to the class frequencies. The data pre-processing was coded in Python, whereas for the logistic regression model the scikit-learn module with the liblinear coordinate descent optimization algorithm was used. Performances of the model were evaluated using the cross-validation confusion matrix as well as by computing precision, sensitivity, specificity, and the Receiver Operating Characteristic (ROC) curve.

### Cell culture and transfection

HEK-Dual™ Null (NF/IL8) cells (Invivogen) cells were cultured in Dulbecco modified Eagle medium (DMEM, Euroclone ECB7501L) supplemented with 10% fetal bovine serum (FBS; Euroclone, ECS0180L), 2 mM L-glutamine and 100 units/ml penicillin-streptomycin at 37°C in an atmosphere of 5% CO2:air. Transfections were performed with 1 µg DNA plasmid using lipofectamine LTX (Life Technologies, 15338500). The cells were seeded to be 70% to 80% confluent, then DNA was diluted in DMEM with 10 mM HEPES. Lipofectamine LTX was next added to the complex (5 µl) to allow creation of complexes (30 min at RT). Ultimately, DNA-lipid complexes were added to cells. Bafilomycin A1 (Santa Cruz Biotechnology, sc-201550). Human primary fibroblasts were obtained from the Genetic Biobank of Siena. Fibroblasts were cultured in Dulbecco’s Modified Eagle’s medium (DMEM) (Biochrom GmbH, Berlin, Germany) supplemented with 10% Fetal Bovine Serum (FBS) (Carlo Erba), 2% L-glutamine (Carlo Erba) and 1% antibiotics (Penicillin/Streptomycin) (Carlo Erba), according to standard protocols, and routinely passed 1:2 with Trypsin/EDTA (0.05%) solution (Irvine Scientific Santa Ana, California, US).

### Immunofluorescence (IF)

Cells were fixed with 4% paraformaldehyde in PBS for 20 min, washed with phosphate-buffered saline (PBS; Oxoid, BR0014G) and then permeabilized with digitonin solution (Life Technologies, BN2006) for 20 minutes. Then, the cells were washed three times in PBS. Permeabilized cells were incubated with anti-LC3B primary antibody (MBL, M152-3) for 1 h, washed three times with PBS, and then incubated with anti-mouse Alexa Fluor 488-conjugated secondary antibody (Life Technologies, A21202); subsequently cells were washed three times with PBS. Nuclei were stained with a solution of 6 µM of 4’,6-diamidino-2-phenylindole (DAPI; Sigma Aldrich, D9542) in PBS for 10 min. Coverslips were mounted in fluorescence mounting medium (Dako, S3023). Samples were visualized on a TSC SP5 confocal microscope (Leica Microsystems, 5100000750) installed on an inverted LEICA DMI 6000CS (Leica Microsystems, 10741320) microscope and equipped with an oil immersion PlanApo 63× 1.4 NA objective. Images were acquired using the LAS AF acquisition software (Leica Microsystems, 10210).

### Dot count and statistical analysis for autophagy

For the LC3B-positive dot count, we performed intensitometric analysis of fluorescence using the Quantitation Module of Volocity software (PerkinElmer Life Science). Dot count was subjected to statistical analysis. Measures were obtained by analyzing at least 400 cells/sample from 3 different experiments. Significance (P value) was assessed by Student’s t test, using GraphPad Prism6 software. Asterisks were attributed for the following significance values: P > 0.05 (ns), P < 0.05 (*), P < 0.01 (**), P < 0.001 (***).

### Real time qPCR analysis of TNF-α expression

Total RNA was isolated using the RNAeasy Mini Kit (Quiagen, Hilden, Germany) according to the manufacturer’s instructions. cDNA synthesis was performed using the Maxima First Strand cDNA Synthesis Kit (Life Technologies, CA, USA). Neo synthetized cDNA was used to perform Real Time PCR using the PowerUp Sybr Green (Life Technologies, CA, USA). Following primers were used TNFα Fw CTATCTGGGAGGGGTCTTCC; TNFα Rw GGTTGAGGGTGTCTGAAGGA; HPRT1 Fw GTCTTGCTCGAGATGTGATG and HPRT1 Rw GTAATCCAGCAGGTCAGCAA. Target transcripts were analyzed with the QuantStudio 7 System (Applied Biosystems, CA, USA). The comparative threshold cycle (Ct) method was used for quantification analysis. The Ct values of each gene were normalized to the Ct value of HPRT1. The gene expression levels were evaluated by the fold change using the equation 2-ΔΔCt.

### *HLA* sequencing

*HLA-class I* and *II* genes were targeted for DNA sequencing using a biotinylated DNA probe-based capture method [PMID: 27486779], with modifications as follows. Genomic DNA (500ng from each sample) was fragmented enzymatically using the NEBNext Ultra ii FS module (New England Biolabs, Boston, MA). Individual samples were labeled uniquely using 3ul of 15uM custom dual-index adapters (Integrated DNA Technologies, Coralville, IA) and the NEB ligation module. Post ligation cleanup was based on the Kapa Hyper Prep protocol (Kapa Biosystems, Wilmington, MA) and followed by dual size selection. Paired ends of 250bp each were sequenced using a NovaSeq instrument and SP Reagent Kit (Illumina Inc, San Diego, CA). *HLA* alleles were determined from the sequence data using the consensus from three algorithms: NGSengine 2.10.0 (GenDX, Utrecht, the Netherlands), HLA Twin (Omixon Biocomputing Ltd. Budapest, Hungary) and HLA*LA [PMID: 30942877].

## Data Availability

The data and samples referenced here are housed in the GEN-COVID Patient Registry and the GEN-COVID Biobank and are available for consultation. You may contact the corresponding author, Prof. Alessandra Renieri.

http://nigdb.cineca.it

https://sites.google.com/dbm.unisi.it/gen-covid

## ACKNOWLEDGEMENTS

This study is part of the GEN-COVID Multicenter Study, https://sites.google.com/dbm.unisi.it/gen-covid, the Italian multicenter study aimed at identifying the COVID-19 host genetic bases. Specimens were provided by the COVID-19 Biobank of Siena, which is part of the Genetic Biobank of Siena, member of BBMRI-IT, of Telethon Network of Genetic Biobanks (project no. GTB18001), of EuroBioBank, and of RD-Connect. We thank the CINECA consortium for providing computational resources and the Network for Italian Genomes (NIG) http://www.nig.cineca.it for its support. We thank private donors for the support provided to A.R. (Department of Medical Biotechnologies, University of Siena) for the COVID-19 host genetics research project (D.L n.18 of March 17, 2020). PJN was supported by a grant from fastgrants.org. We also thank the COVID-19 Host Genetics Initiative (https://www.covid19hg.org/) and MIUR project “Dipartimenti di Eccellenza 2018-2020” to the Department of Medical Biotechnologies University of Siena, Italy. We thanks Dr. Margherita Leonardi for the experimental contribution in the autophagy data analysis. TDJF has received the Post-Doctoral fellowship from Conselho Nacional de Desenvolvimento Científico e Tecnológico (CNPq) of Brazil.

## ETHICS APPROVAL

The GEN-COVID study was approved by the University Hospital of Siena Ethical Review Board (Protocol n. 16929, dated March 16, 2020).

## AUTHOR CONTRIBUTIONS

SC, CF and IM were in charge of biological samples’ collection and biobanking. CF and SD were in charge of DNA isolations from peripheral blood samples and carried out the sequencing experiments. MB, MP, FF and FMo and FMa were in charge of clinical data collection. EB, NP, and SF performed bioinformatics and statistical analyses. MLC, LDS, GB and DA prepared Figures and Tables. MB, FF, AR, FM, AMI, MUM, CB and AA performed analysis/interpretation of clinical data. MAV and FC were in charge of functional analysis for TLR3 L142F activity in HEK cells. MVC and OS performed structural biology analysis on protein models. FI performed autophagy experiments. SA provided TLR3 plasmids. SC, MUM, IM, AR, SM wrote and edited the manuscript. MUM, AR, IM, AMI, AA, MC, EF and FM designed the study. TDJf and PJN performed HLA sequencing. AA, SC and IM analysed HLA data. All authors have reviewed and approved the manuscript.

## GEN-COVID Multicenter Study (https://sites.google.com/dbm.unisi.it/gen-covid)

Mirella Bruttini^1,2,3^, Rossella Tita^3^, Sara Amitrano^3^, Anna Maria Pinto^3^, Maria Antonietta Mencarelli^3^, Caterina Lo Rizzo^3^, Valentina Perticaroli^1,2,3^, Massimiliano Fabbiani^9^, Barbara Rossetti^9^, Giacomo Zanelli^2,9^, Elena Bargagli^10^, Laura Bergantini^10^, Miriana D’Alessandro^10^, Paolo Cameli^10^, David Bennett^10^, Federico Anedda^11^, Simona Marcantonio^11^, Sabino Scolletta^11^, Federico Franchi^11^, Maria Antonietta Mazzei^12^, Susanna Guerrini^12^, Edoardo Conticini^13^, Luca Cantarini^13^, Bruno Frediani^13^, Danilo Tacconi^14^, Chiara Spertilli^14^, Marco Feri^15^, Alice Donati^15^, Raffaele Scala^16^, Luca Guidelli^16^, Genni Spargi^17^, Marta Corridi^17^, Cesira Nencioni^18^, Leonardo Croci^18^, Gian Piero Caldarelli^19^, Maurizio Spagnesi^20^, Paolo Piacentini^20^, Maria Bandini^20^, Elena Desanctis^20^, Silvia Cappelli^20^, Anna Canaccini^21^, Agnese Verzuri^21^, Valentina Anemoli^21^, Agostino Ognibene^22^, Alessandro Pancrazi^22^, Maria Lorubbio^22^, Massimo Vaghi^23^, Antonella D’Arminio Monforte^24^, Esther Merlini^24^, Federica Gaia Miraglia^24^, Raffaele Bruno^25,26^, Marco Vecchia^25^, Serena Ludovisi^25,26^, Massimo Girardis^27^, Sophie Venturelli^27^, Marco Sita^27^, Andrea Cossarizza^28^, Andrea Antinori^29^, Alessandra Vergori^29^, Arianna Emiliozzi^29^, Stefano Rusconi^30,31^, Matteo Siano^31^, Arianna Gabrieli^31^, Agostino Riva^30,31^, Daniela Francisci^32,33^, Elisabetta Schiaroli^32^, Francesco Paciosi^32^, Pier Giorgio Scotton^34^, Francesca Andretta^34^, Sandro Panese^35^, Renzo Scaggiante^36^, Francesca Gatti^36^, Saverio Giuseppe Parisi^37^, Melania degli Antoni^38^, Isabella Zanella^39,40^, Matteo Della Monica^41^, Carmelo Piscopo^41^, Mario Capasso^42,43,44^, Roberta Russo^42,43^, Immacolata Andolfo^42,43^, Achille Iolascon^42,43^, Giuseppe Fiorentino^45^, Massimo Carella^46^, Marco Castori^46^, Filippo Aucella^47^, Pamela Raggi^48^, Carmen Marciano^48^, Rita Perna^48^, Matteo Bassetti^49,50^, Antonio Di Biagio^50^, Maurizio Sanguinetti^51,52^, Luca Masucci^51,52^, Serafina Valente^53^, Marco Mandalà^54^, Alessia Giorli^54^, Lorenzo Salerni^54^, Patrizia Zucchi^55^, Pierpaolo Parravicini^55^, Elisabetta Menatti^56^, Stefano Baratti^57^, Tullio Trotta^58^, Ferdinando Giannattasio^58^, Gabriella Coiro^58^, Fabio Lena^59^, Domenico A. Coviello^60^, Cristina Mussini^61^, Giancarlo Bosio^62^, Enrico Martinelli^62^, Sandro Mancarella^63^, Luisa Tavecchia^63^, Mary Ann Belli^63^, Lia Crotti^64,65,66,67^, Gianfranco Parati^64,65^, Marco Gori^68,69^, Maurizio Sanarico^70^, Stefano Ceri^71^, Pietro Pinoli^71^, Francesco Raimondi^72^, Filippo Biscarini^73^, Alessandra Stella^73^, Marco Rizzi^74^, Franco Maggiolo^74^, Diego Ripamonti^74^, Claudia Suardi^75^, Tiziana Bachetti^76^, Maria Teresa La Rovere^77^, Simona Sarzi-Braga^78^, Maurizio Bussotti^79^, Mattia Bergomi^80^, Katia Capitani^2,81^, Kristina Zguro^2^, Simona Dei^82^, Sabrina Ravaglia^83^, Rosangela Artuso^84^, Antonio Perrella^85^, Francesco Bianchi^2,85^, Giuseppe Merla^42,86^, Gabriella Maria Squeo^86^.

9. Dept of Specialized and Internal Medicine, Tropical and Infectious Diseases Unit, Azienda Ospedaliera Universitaria Senese, Siena, Italy

10. Unit of Respiratory Diseases and Lung Transplantation, Department of Internal and Specialist Medicine, University of Siena

11. Dept of Emergency and Urgency, Medicine, Surgery and Neurosciences, Unit of Intensive Care Medicine, Siena University Hospital, Italy

12. Department of Medical, Surgical and Neurosciences and Radiological Sciences, Unit of Diagnostic Imaging, University of Siena

13. Rheumatology Unit, Department of Medicine, Surgery and Neurosciences, University of Siena, Policlinico Le Scotte, Italy

14. Department of Specialized and Internal Medicine, Infectious Diseases Unit, San Donato Hospital Arezzo, Italy

15. Dept of Emergency, Anesthesia Unit, San Donato Hospital, Arezzo, Italy

16. Department of Specialized and Internal Medicine, Pneumology Unit and UTIP, San Donato Hospital, Arezzo, Italy

17. Department of Emergency, Anesthesia Unit, Misericordia Hospital, Grosseto, Italy

18. Department of Specialized and Internal Medicine, Infectious Diseases Unit, Misericordia Hospital, Grosseto, Italy

19. Laboratory Medicine Department, Misericordia Hospital, Grosseto, Italy

20. Department of Preventive Medicine, Azienda USL Toscana Sud Est, Italy

21. Territorial Scientific Technician Department, Azienda USL Toscana Sud Est, Italy

22. Laboratory Medicine Department, San Donato Hospital, Arezzo, Italy

23. Chirurgia Vascolare, Ospedale Maggiore di Crema, Italy

24. Department of Health Sciences, Clinic of Infectious Diseases, ASST Santi Paolo e Carlo, University of Milan, Italy

25. Division of Infectious Diseases and Immunology, Fondazione IRCCS Policlinico San Matteo, Pavia, Italy

26. Department of Internal Medicine and Therapeutics, University of Pavia, Italy

27. Department of Anesthesia and Intensive Care, University of Modena and Reggio Emilia, Modena, Italy

28. Department of Medical and Surgical Sciences for Children and Adults, University of Modena and Reggio Emilia, Modena, Italy

29. HIV/AIDS Department, National Institute for Infectious Diseases, IRCCS, Lazzaro Spallanzani, Rome, Italy

30. III Infectious Diseases Unit, ASST-FBF-Sacco, Milan, Italy

31. Department of Biomedical and Clinical Sciences Luigi Sacco, University of Milan, Milan, Italy

32. Infectious Diseases Clinic, Department of Medicine, Azienda Ospedaliera di Perugia and University of Perugia, Santa Maria Hospital, Perugia, Italy

33. Infectious Diseases Clinic, “Santa Maria” Hospital, University of Perugia, Perugia, Italy

34. Department of Infectious Diseases, Treviso Hospital, Local Health Unit 2 Marca Trevigiana, Treviso, Italy

35. Clinical Infectious Diseases, Mestre Hospital, Venezia, Italy.

36. Infectious Diseases Clinic, ULSS1, Belluno, Italy

37. Department of Molecular Medicine, University of Padova, Italy

38. Department of Infectious and Tropical Diseases, University of Brescia and ASST Spedali Civili Hospital, Brescia, Italy

39. Department of Molecular and Translational Medicine, University of Brescia, Italy;

40. Clinical Chemistry Laboratory, Cytogenetics and Molecular Genetics Section, Diagnostic Department, ASST Spedali Civili di Brescia, Italy

41. Medical Genetics and Laboratory of Medical Genetics Unit, A.O.R.N. “Antonio Cardarelli”, Naples, Italy

42. Department of Molecular Medicine and Medical Biotechnology, University of Naples Federico II, Naples, Italy

43. CEINGE Biotecnologie Avanzate, Naples, Italy

44. IRCCS SDN, Naples, Italy

45. Unit of Respiratory Physiopathology, AORN dei Colli, Monaldi Hospital, Naples, Italy

46. Division of Medical Genetics, Fondazione IRCCS Casa Sollievo della Sofferenza Hospital, San Giovanni Rotondo, Italy

47. Department of Medical Sciences, Fondazione IRCCS Casa Sollievo della Sofferenza Hospital, San Giovanni Rotondo, Italy

48. Clinical Trial Office, Fondazione IRCCS Casa Sollievo della Sofferenza Hospital, San Giovanni Rotondo, Italy

49. Department of Health Sciences, University of Genova, Genova, Italy

50. Infectious Diseases Clinic, Policlinico San Martino Hospital, IRCCS for Cancer Research Genova, Italy

51. Microbiology, Fondazione Policlinico Universitario Agostino Gemelli IRCCS, Catholic University of Medicine, Rome, Italy

52. Department of Laboratory Sciences and Infectious Diseases, Fondazione Policlinico Universitario A. Gemelli IRCCS, Rome, Italy

53. Department of Cardiovascular Diseases, University of Siena, Siena, Italy

54. Otolaryngology Unit, University of Siena, Italy

55. Department of Internal Medicine, ASST Valtellina e Alto Lario, Sondrio, Italy

56. Study Coordinator Oncologia Medica e Ufficio Flussi, Sondrio, Italy

57. Department of Infectious and Tropical Diseases, University of Padova, Padova, Italy

58. First Aid Department, Luigi Curto Hospital, Polla, Salerno, Italy

59. Local Health Unit-Pharmaceutical Department of Grosseto, Toscana Sud Est Local Health Unit, Grosseto, Italy

60. U.O.C. Laboratorio di Genetica Umana, IRCCS Istituto G. Gaslini, Genova, Italy.

61. Infectious Diseases Clinics, University of Modena and Reggio Emilia, Modena, Italy.

62. Department of Respiratory Diseases, Azienda Ospedaliera di Cremona, Cremona, Italy

63. U.O.C. Medicina, ASST Nord Milano, Ospedale Bassini, Cinisello Balsamo (MI), Italy

64. Istituto Auxologico Italiano, IRCCS, Department of Cardiovascular, Neural and Metabolic Sciences, San Luca Hospital, Milan, Italy.

65. Department of Medicine and Surgery, University of Milano-Bicocca, Milan, Italy

66. Istituto Auxologico Italiano, IRCCS, Center for Cardiac Arrhythmias of Genetic Origin, Milan, Italy.

67. Istituto Auxologico Italiano, IRCCS, Laboratory of Cardiovascular Genetics, Milan, Italy.

68. University of Siena, DIISM-SAILAB, Siena, Italy

69. University Cote d’Azur, Inria, CNRS, I3S, Maasai

70. Independent Data Scientist, Milan, Italy

71. Department of Electronics, Information and Bioengineering (DEIB), Politecnico di Milano, Milano, Italy

72. Scuola Normale Superiore, Pisa, Italy

73. CNR-Consiglio Nazionale delle Ricerche, Istituto di Biologia e Biotecnologia Agraria (IBBA), Milano, Italy.

74. Unit of Infectious Diseases, ASST Papa Giovanni XXIII Hospital, Bergamo, Italy

75. Fondazione per la ricerca Ospedale di Bergamo, Bergamo, Italy

76. Direzione Scientifica, Istituti Clinici Scientifici Maugeri IRCCS, Pavia, Italy.

77. Istituti Clinici Scientifici Maugeri IRCCS, Department of Cardiology, Institute of Montescano, Pavia, Italy.

78. Istituti Clinici Scientifici Maugeri, IRCCS, Department of Cardiac Rehabilitation, Institute of Tradate (VA), Italy.

79. Cardiac Rehabilitation Unit, Fondazione Salvatore Maugeri, IRCCS, Scientific Institute of Milan, Milan, Italy.

80. Veos Digital, Milan, Italy

81. Core Research Laboratory, ISPRO, Florence, Italy

82. Health Management, Azienda USL Toscana Sudest, Tuscany, Italy

83. IRCCS C. Mondino Foundation, Pavia, Italy

84. Medical Genetics Unit, Meyer Children’s University Hospital, Florence, Italy

85. Department of Medicine, Pneumology Unit, Misericordia Hospital, Grosseto, Italy.

86. Laboratory of Regulatory and Functional Genomics, Fondazione IRCCS Casa Sollievo della Sofferenza, San Giovanni Rotondo (Foggia), Italy

